# From Concept to Code: AI- Powered CODE-ICH Transforming Acute Neurocritical Response for Hemorrhagic Strokes

**DOI:** 10.1101/2025.09.26.25336582

**Authors:** Saif Salman, Rosa Corro, Terri Menser, Devang Sanghavi, Christopher Kramer, Pablo Moreno Franco, WD freeman

**Author notes:** Corresponding Author: William D. Freeman, MD, Department of Neurologic Surgery, Mayo Clinic, 4500 San Pablo Rd, Jacksonville, FL 32224.

## Abstract

**Background:** Intracerebral hemorrhage (ICH) is among the most devastating forms of stroke, characterized by high early mortality and limited time-sensitive treatment protocols compared to ischemic stroke. The absence of standardized emergency response frameworks and the shortcomings of conventional scoring systems highlight the urgent need for innovation in neurocritical care.

**Objective:** This paper introduces and evaluates the CODE-ICH framework, along with two AI-powered tools HEADS-UP and SAHVAI designed to transform acute ICH management through real-time detection, volumetric analysis, and predictive modeling.

**Methods:** We describe the development and implementation of HEADS-UP, a cloud-based AI system for early ICH detection in underserved populations, and SAHVAI, a convolutional neural network–based tool for subarachnoid hemorrhage volume quantification. These tools were integrated into a novel paging and workflow system at a comprehensive stroke center to facilitate ultra-early intervention.

**Results:** SAHVAI achieved 99.8% accuracy in volumetric analysis and provided 2D, 3D, and 4D visualization of hemorrhage progression. HEADS-UP enabled rapid triage and transfer, reducing reliance on subjective interpretation. Together, these tools operationalized the time is brain principle for hemorrhagic stroke and supported proactive, data-driven care in the neuro–intensive care unit (NICU).

**Conclusion:** CODE-ICH, HEADS-UP, and SAHVAI represent a paradigm shift in hemorrhagic stroke care, delivering scalable, explainable, and multimodal AI solutions that enhance clinical decision-making, minimize delays, and promote equitable access to neurocritical care.

## Introduction

Hemorrhagic strokes are among the most devastating conditions encountered in the neuro– intensive care unit (NICU). Despite advances in medical management and surgical interventions, outcomes remain poor, with 30-day mortality rates approaching 40% (1). Acute care for intracerebral hemorrhage (ICH) still lags behind that for ischemic stroke, with no widely adopted time-based emergency protocols. This disparity is concerning given the downstream complications that often culminate in death. There is an urgent need to address this gap and ensure timely, standardized care for ICH patients (2,3).

The CODE-ICH framework positions ICH as an ultra-emergency, aligning it with the “time is brain” principle well established in ischemic stroke (4–7). Brott et al. demonstrated the heterogeneous trajectories of hemorrhagic volume progression, showing that early hematoma growth reflects an accelerated rate of neuronal destruction. They further emphasized that neuronal loss during this rapid expansion phase far exceeds that of ischemic stroke, underscoring the relevance of the “time is brain” paradigm in ICH (8). Larger static hematomas (33–45 mL) and rapidly expanding bleeds (>100 mL within hours) can result in catastrophic neuronal loss, highlighting the need for ultra-early intervention (Figure 1) (8).

**Figure 1:**
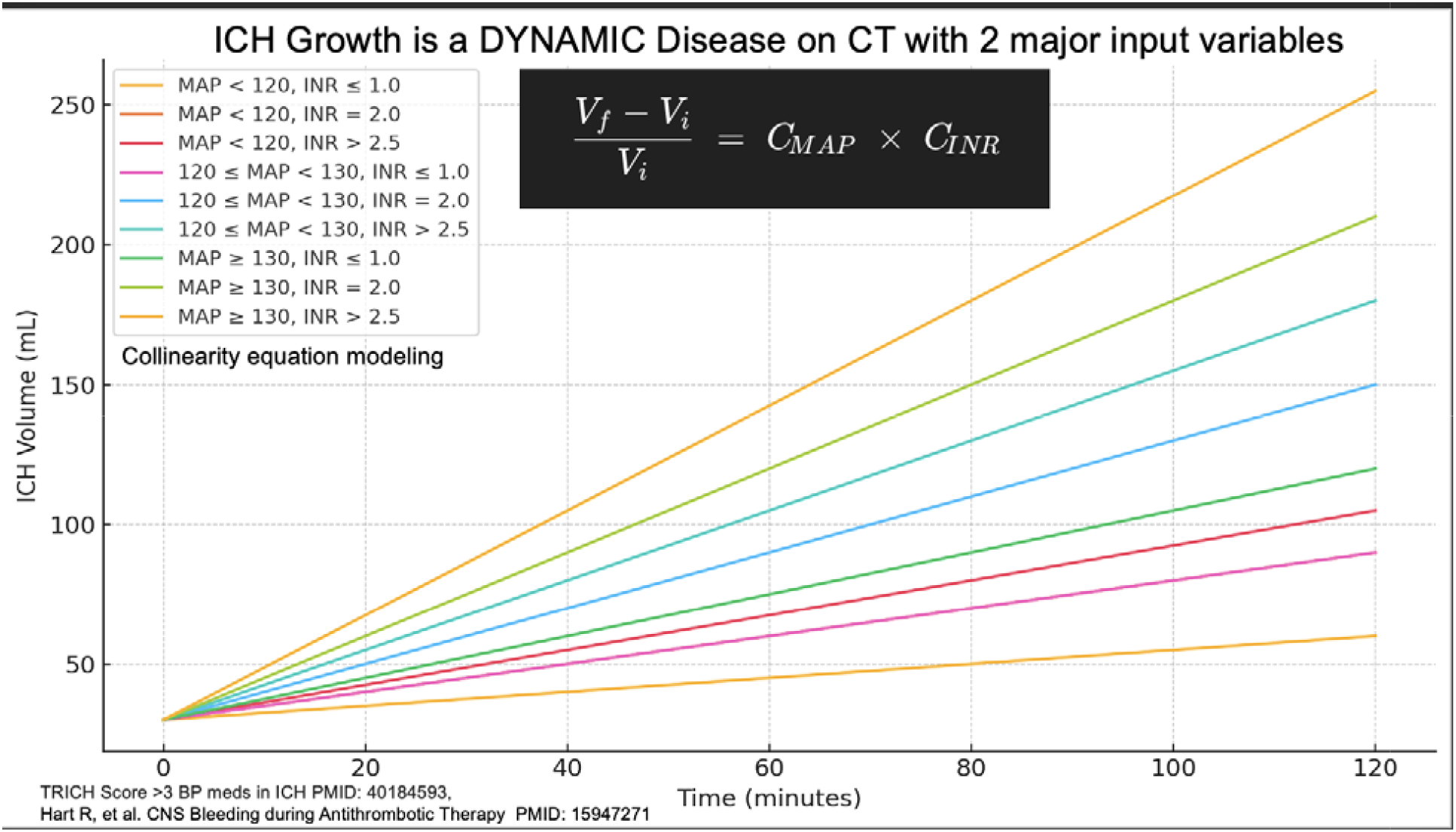
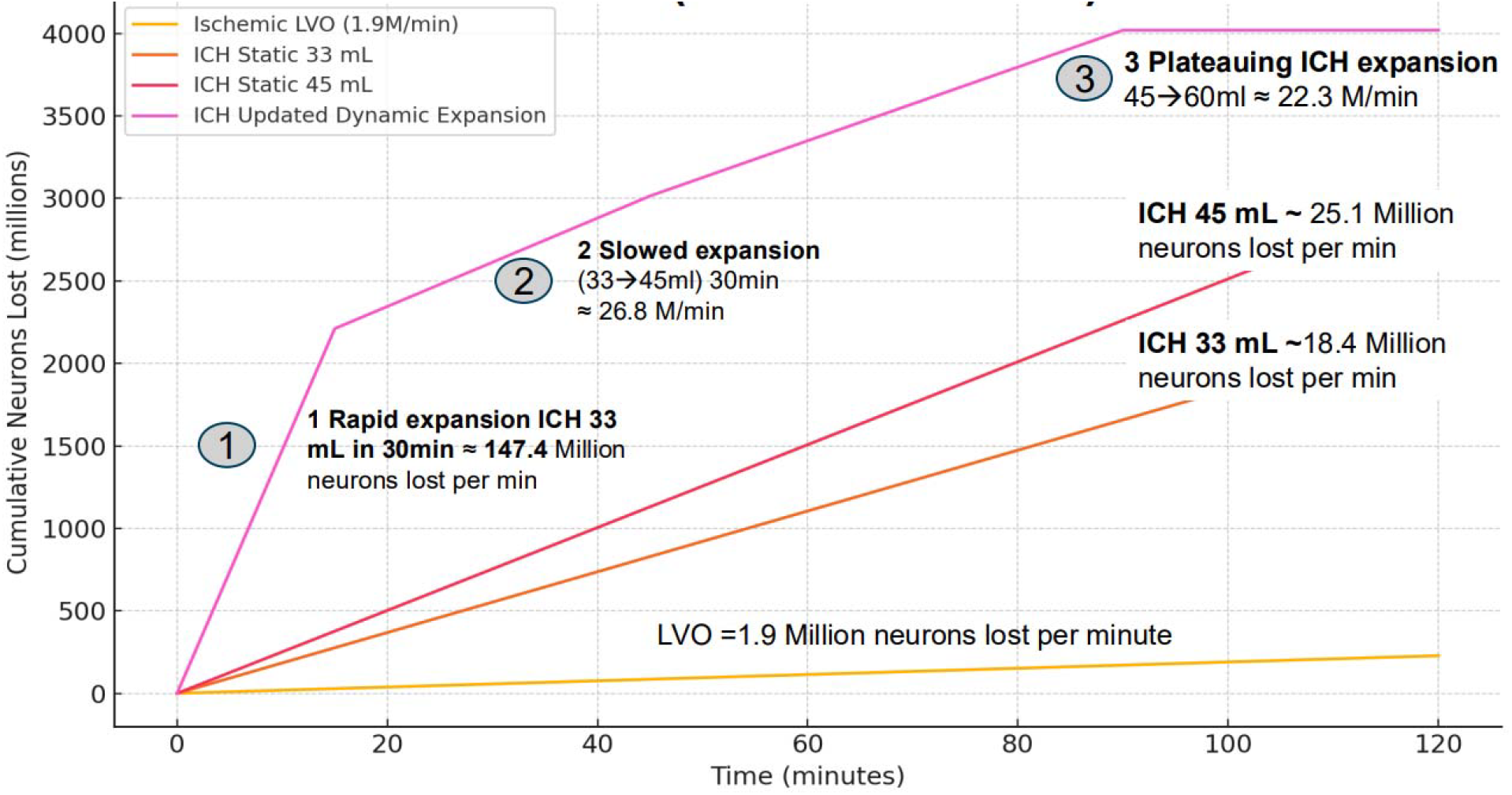
A) Conceptual model of ICH as a dynamic and evolving process, the graph illustrates the synergistic influence of blood pressure (MAP) and coagulation profile (INR) on hematomal growth over time. It shows different trajectories of hemorrhagic expansion based on patient stratification into groups based on MAP and INR B) Estimated neuronal loss rates. Ischemic large-vessel occlusion results in ∼1.9 million neurons/min, while ICH results in greater losses, exceeding 25 million neurons/min in static hematomas and more than 147 million neurons/min during rapid expansion

Although guidelines such as INTERACT-3 emphasize bundled care, their implementation is limited by NICU workforce shortages and delays in multimodal neuromonitoring interpretation (1,9). Conventional scales such as the modified Fisher scale (mFS) and the ICH score lack predictive power, are subject to inter-rater variability, and cannot adapt dynamically. Thus, AI integration into NICU workflows becomes essential when traditional scoring systems fall short or staffing limitations persist (10).

Neurointensivists face an overwhelming influx of data, including invasive monitoring, imaging, and continuous hemodynamic recordings, all requiring rapid and rigorous interventions. Artificial intelligence (AI) can streamline this influx into unified workflows. Current applications include automated hemorrhage detection and quantification, aneurysm risk stratification, and seizure detection as a complication of hemorrhage. AI also enables identification of subtle radiologic and physiologic markers invisible to the human eye, thereby improving early detection of hemorrhage expansion and vasospasm, particularly for less experienced clinicians (1,10–13).

Beyond diagnostics, predictive AI models are transitioning from theoretical frameworks to practical clinical integration. These models create “virtual NICU” environments to test management strategies by incorporating multimodal data—bedside examination, clinical scores, laboratory results, and imaging. This approach transforms care from reactive to proactive by predicting outcomes, anticipating secondary injuries, and supporting timely interventions. Natural language processing (NLP) and large language models (LLMs) further expand this horizon by reducing documentation burdens, organizing complex data streams, and supporting clinical decision-making (11).

Multiple studies have validated automated detection, segmentation, and quantification of hemorrhagic strokes (14–16), accelerated by the availability of the 2019 RSNA dataset (17). Additional work has focused on predicting hematoma expansion—a fatal event within the first 6 hours—along with delayed cerebral ischemia (DCI) and mortality (18–22). Collectively, these studies confirm AI’s viability in detection, quantification, and outcome prediction.

Building on the CODE-ICH framework and the work of Brott et al., expedited neuroimaging and AI-based hemorrhage quantification hold promise for significantly improving outcomes, particularly during the rapid expansion phase (8). Substantial neuronal loss occurs during the “golden hour” of imaging interpretation and report generation. AI tools such as SAHVAI address this gap by providing immediate volumetric and prognostic outputs, embedding CODE-ICH principles directly into NICU workflows.

### HEADS-UP and SAHVAI: Multimodal AI for Real-Time Neurocritical Care

The current state of subarachnoid hemorrhage (SAH) care is hampered by delays in detection and interpretation, inter-rater variability, and reliance on subjective clinical scales. The modified Fisher scale (mFS), widely used for DCI prediction, yields an area under the ROC curve of only 0.67, reflecting major limitations (23). Manual segmentation of subarachnoid hemorrhage volume (SAHV) is considered the gold standard for quantification but is time-consuming, labor-intensive, and impractical in emergencies (24).

On December 19, 2022, the Mayo Clinic Comprehensive Stroke Center (CSC) launched the “ICH Phases” paging system, designed to accelerate team response in ICH cases. The system incorporates AI, fosters holistic engagement of multidisciplinary teams, enhances communication, and establishes benchmarks aligned with AHA/ASA guidelines (Figure 2). AI integration during Phase I (detection) directs SAHV quantification and outcome prediction, enabling timely intervention in subsequent phases.

**Figure 2:**
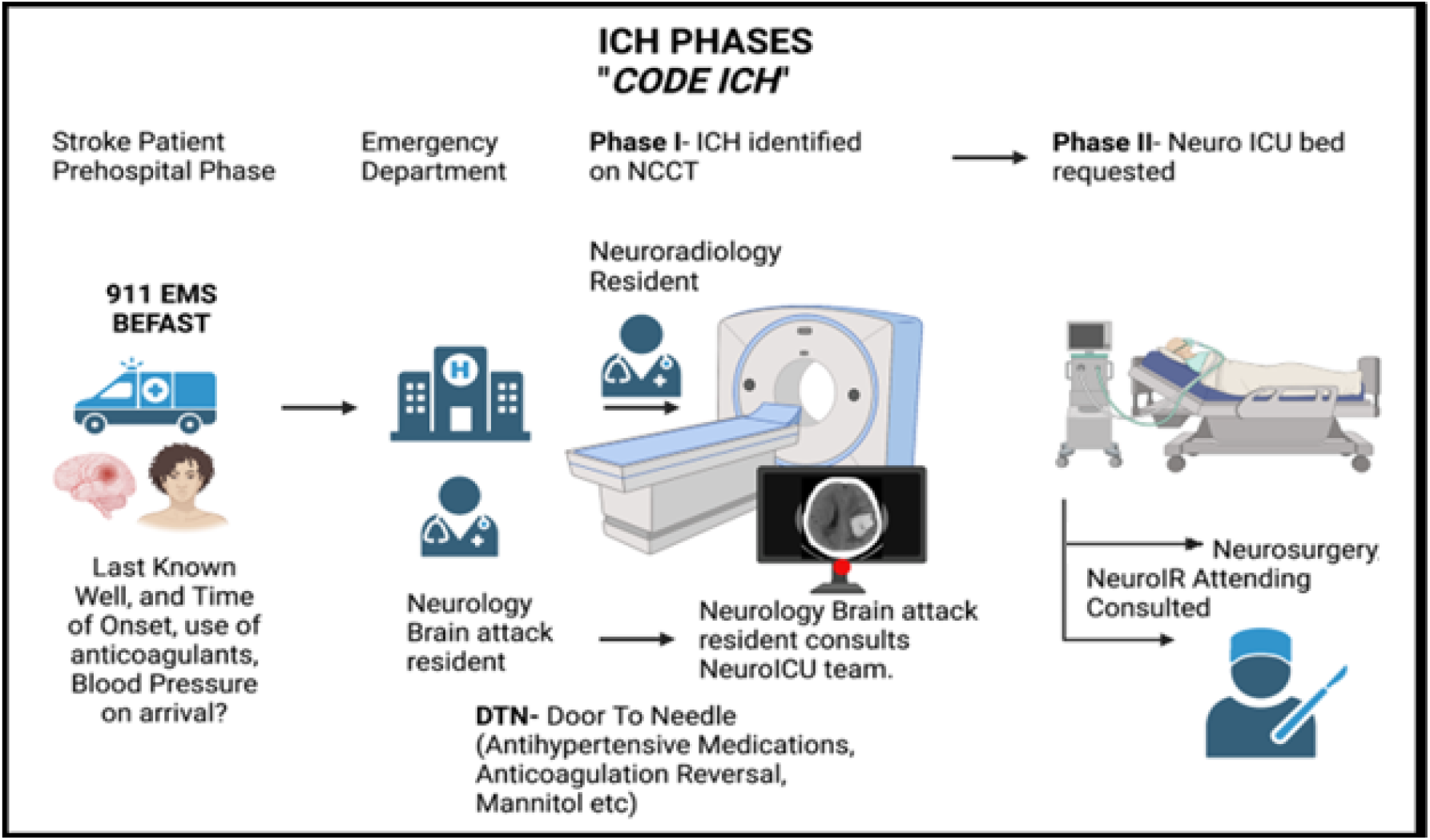
Visual Flow Diagram for “ICH CARE” Phases of communication system launched by the Mayo Clinic Comprehensive Stroke Center (CSC)

**Figure 3:**
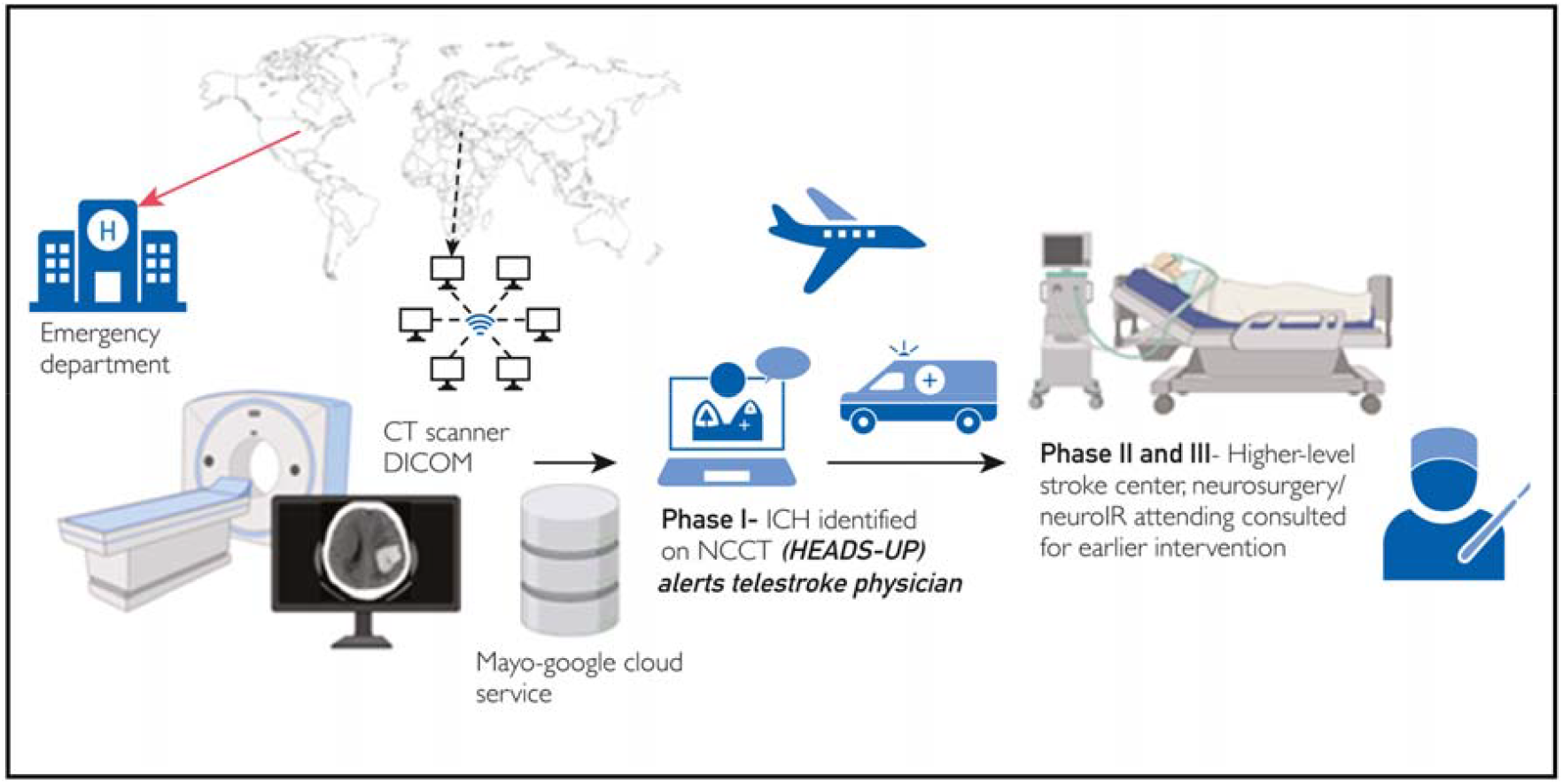
Visual Flow Diagram for HEADS-UP accelerating the detection phase of ICH.

The Hemorrhage Evaluation and Detector System in Underserved Populations (HEADS-UP) reshapes acute care by leveraging cloud-based AI detection. Non-contrast CT (NCCT) scans acquired in the emergency department (ED) are transmitted as DICOM files to a secure Mayo–Google Cloud platform, where AI algorithms promptly identify ICH and alert on-call physicians. This Phase I recognition not only saves time but also reduces reliance on subjective human interpretation. Phases II and III then facilitate expedited transfer to higher-level stroke centers, bridging geographic disparities in access to expertise and improving outcomes (Figure 2) (25).

Subarachnoid Hemorrhage Volumetric Analysis via AI (SAHVAI) uses convolutional neural networks trained on NCCT scans to quantify SAHV in under 10 seconds with 99.8% accuracy. SAHV >10 mL at admission triples the odds of poor outcome (mRS 4–6) and doubles the risk of complications such as DCI. By enabling immediate quantification in the ED, SAHVAI operationalizes CODE-ICH and the “time is brain” principle for hemorrhagic stroke (Figure 4a).

**Figure 4:**
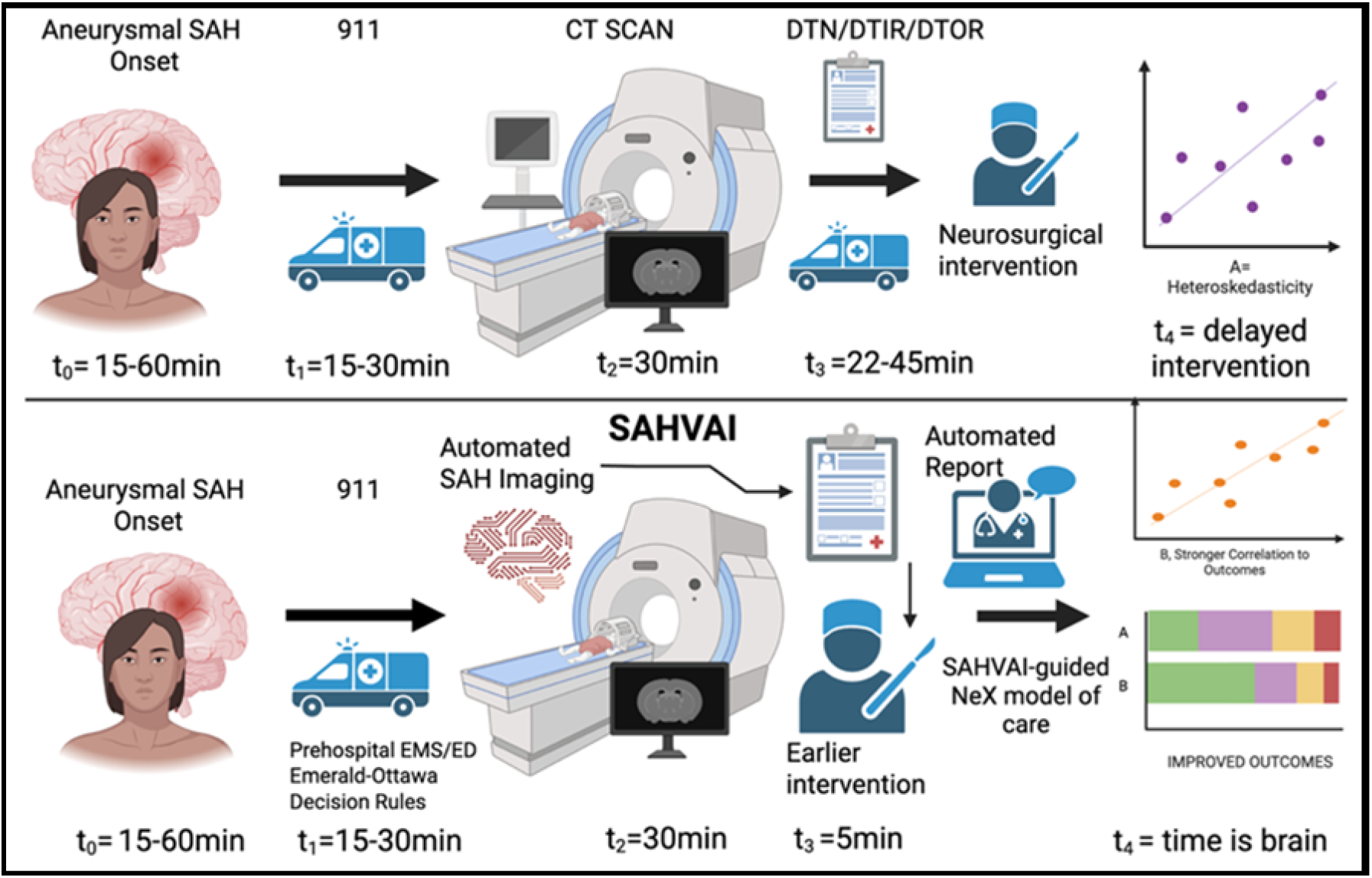
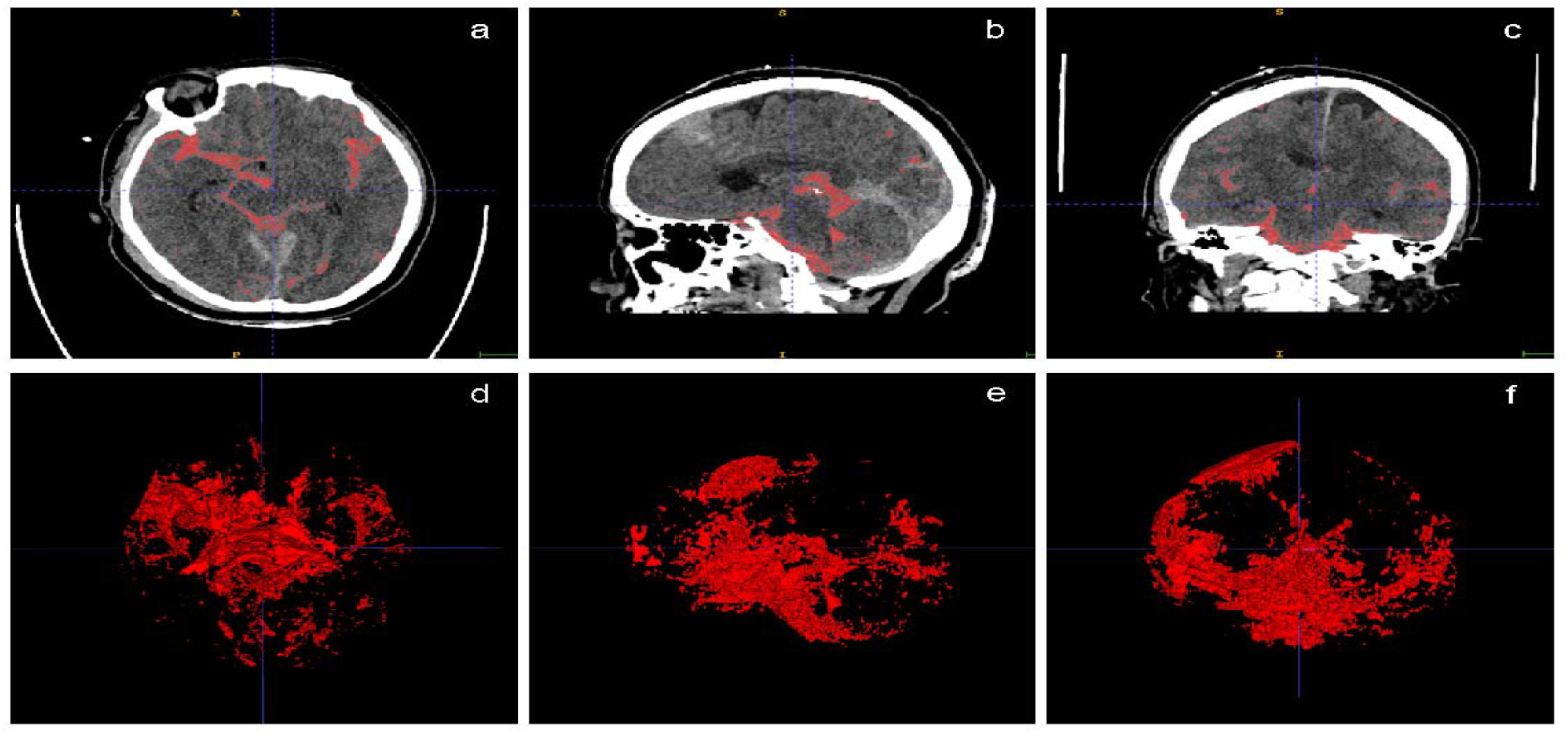
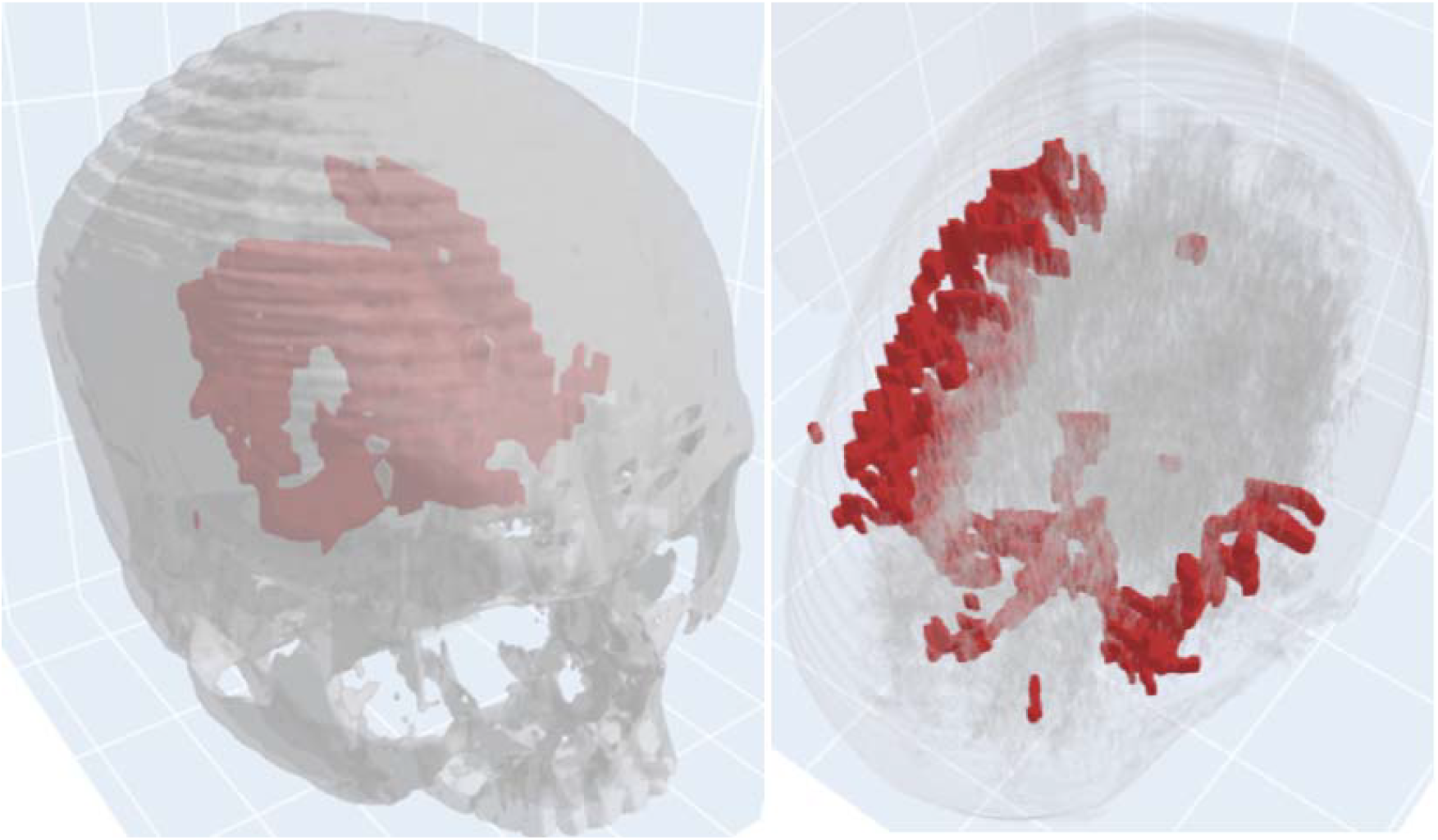
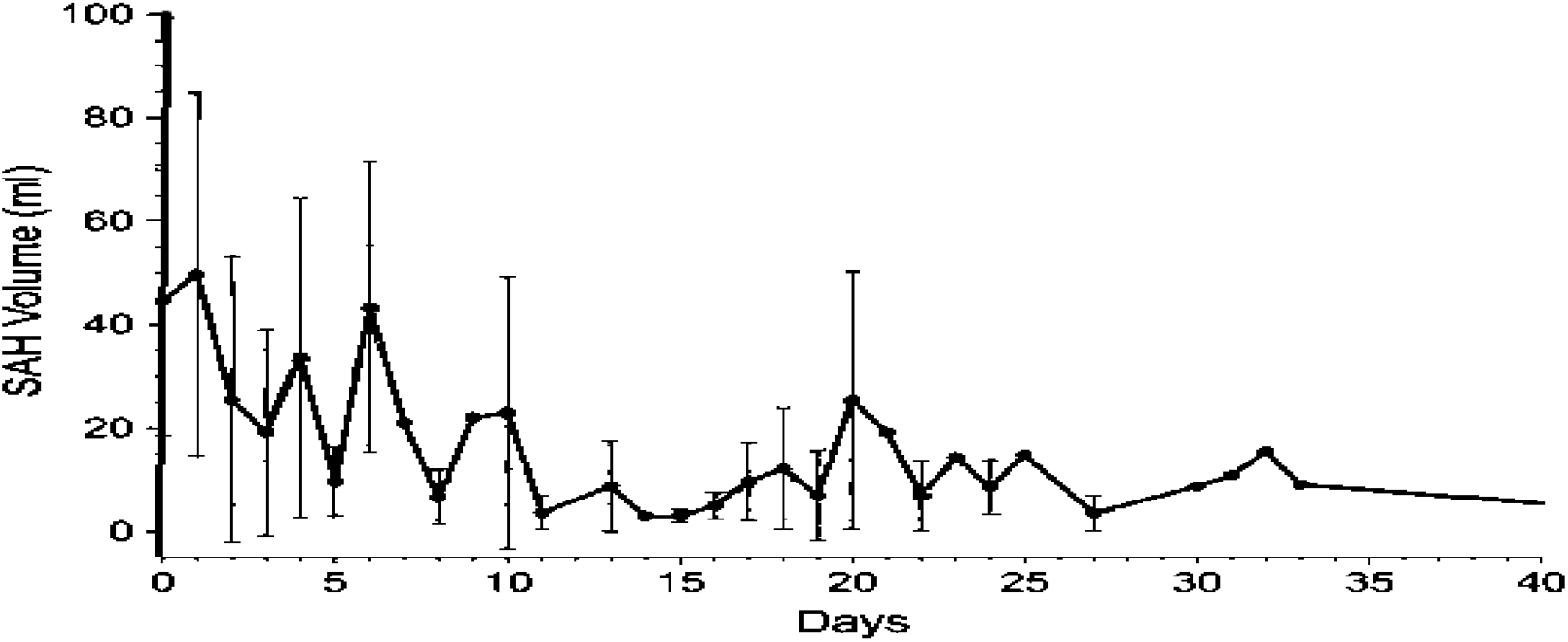
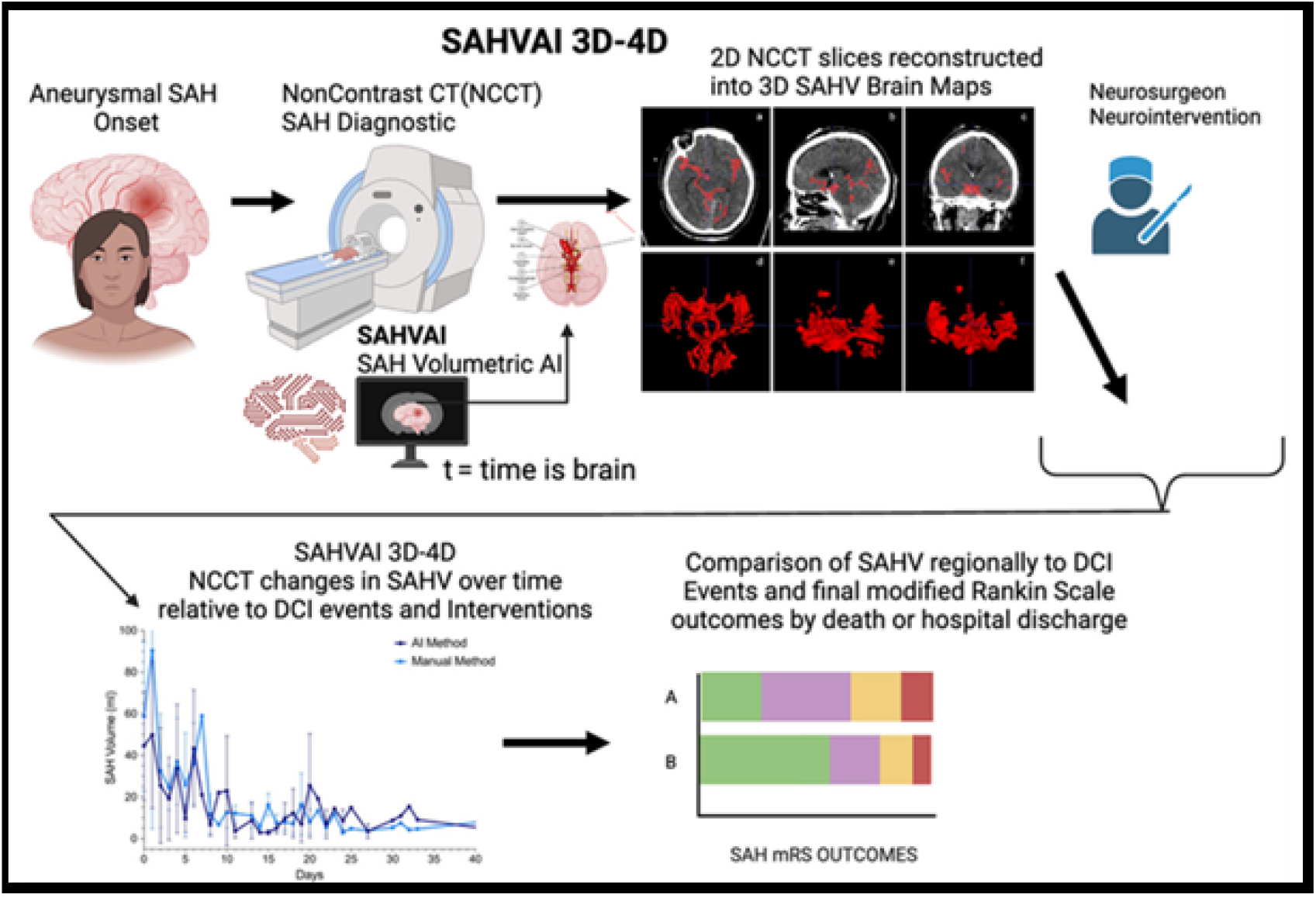
A) Representation of the framework wherein SAHVAI optimizes quantification of SAHV and allowing interventions in a timely manner. B) SAHVAI-2D and SAHVAI-3D brain maps on admission. Segmentation of 5 cisternal spaces; red indicates blood. Planes (2D): (A) axial; (B) sagittal; (C) coronal. View (3D): (D) axial; (E) sagittal; (F) coronal. 2D indicates 2-dimensional; 3D, 3-dimensional C) Anterior and superior views of the SAHVAI-3D brain maps **D)** SAHVAI-4D: Plot of the mean (SD) SAHV for study cohort (N = 10) over time. E) **SAHV, SAHVAI, SAHVAI-3D**, Upper image showing SAHVAI workflow with patient gets non-contrast CT head image on CT table, image loaded into PACS- DICOM, where DICOM is ingested into SAHVAI model for rapid SAHV output in 2D slices and SAHV (total volume), and later can be output into a 3D Image (SAHVAI-3D). These 3D images can be used by clinicians to see brain-regions at highest risk with thickest SAH blood accumulation for future DCI “at-risk” brain regions. Bottom image: End-result of SAHVAI in 2-dimensional (a, b, c) and 3-dimensional (d, e, f) subarachnoid hemorrhage volume, SAHV-3D map

The strength of SAHVAI lies not only in SAHV quantification but also in its multimodal platform capabilities. Continuous SAHV quantification and 3D SAHV map generation (Figures 4b and 4c) visualize high-risk territories for DCI and guide intervention strategies. By extending to 4D analysis, SAHVAI tracks SAHV over time (Figure 4d), capturing the dynamic progression of hemorrhage and correlating it with interventions and complications such as DCI (Figure 4e).

Beyond imaging, SAHVAI integrates electronic medical record (EMR) data including intracranial pressure (ICP), cerebral perfusion pressure (CPP), electroencephalography (EEG), and transcranial Doppler (TCD) ultrasound. This integration facilitates early detection of complications such as DCI, hydrocephalus, and seizures (Figure 5a). Continuous SAHV measurement also defines interventional thresholds: for example, SAHV >10 mL is associated with the need for CSF diversion and ultra-early blood pressure stabilization. Combined 3D and temporal (4D) analyses predict DCI risk and identify areas vulnerable to ischemic injury. Collectively, SAHVAI functions as a holistic, multimodal system producing timely outputs that support proactive neurocritical care (Figure 5b).

**Figure 5.**
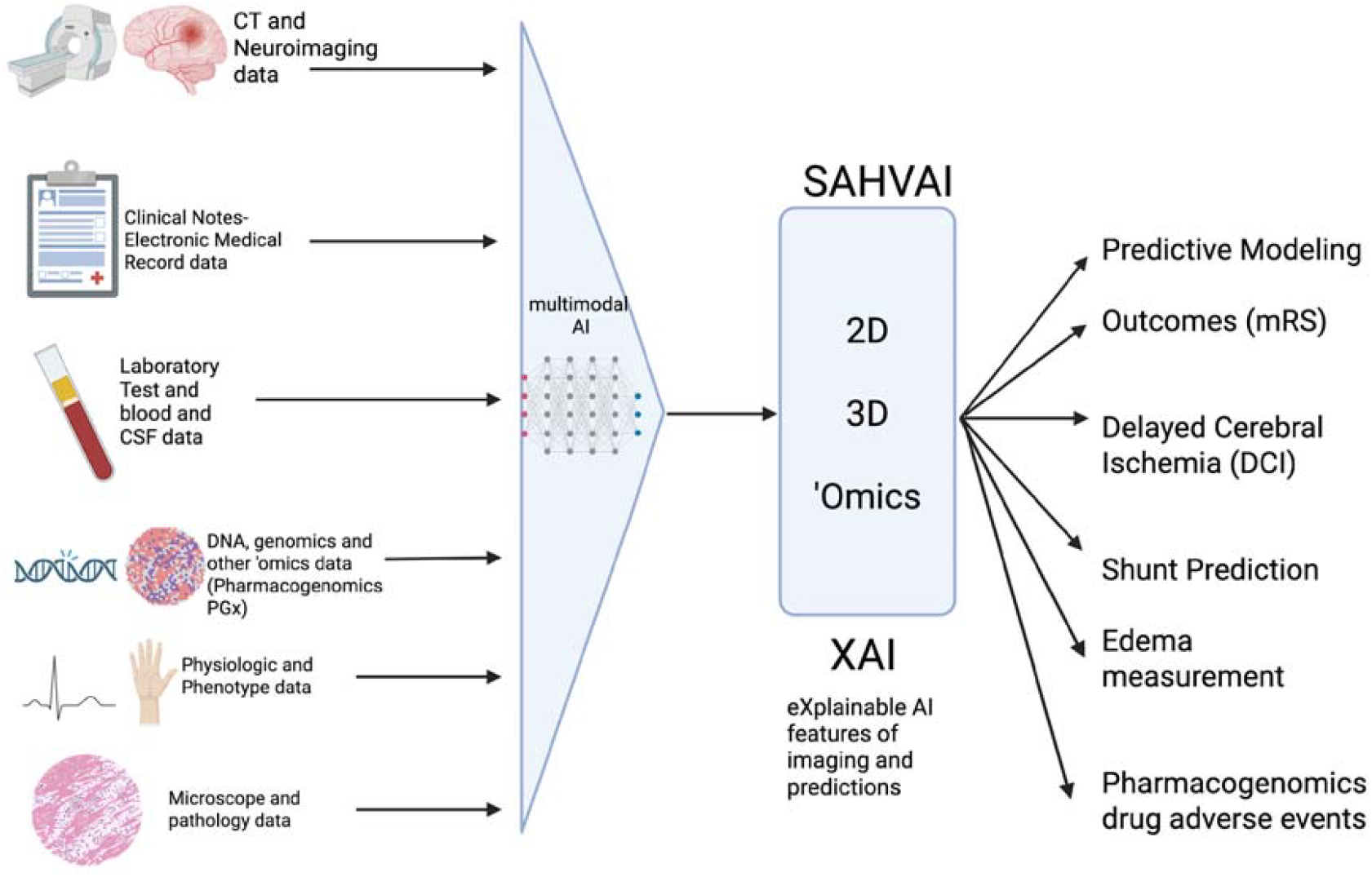
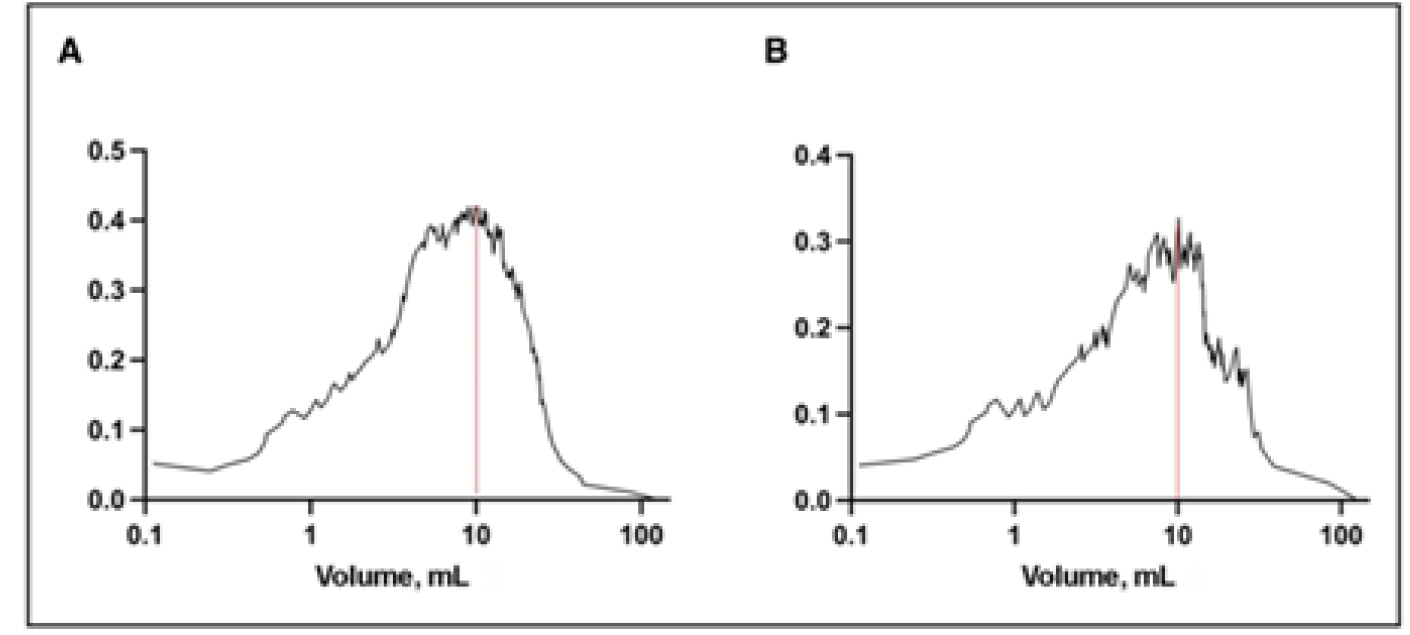
a) Multimodality in SAHVAI incorporating NCCT, EMR, and lab results to predict outcomes. b) subarachnoid hemorrhage volume (SAHV) Youden index (J = sensitivity + specificity – 1) for each value of predictor variables. Dashed lines indicate maximum Youden index. A, JABC-SAHV-outcome=10.086 mL. B, JABC-SAHV-DCI=10.117 mL. DCI indicates delayed cerebral ischemia.

### Explainability: Approach to AI and XAI Imaging (SAHDAI)

The first approach involves creating **SAH detection capabilities** within any DICOM image **(HEADS-UP)**, followed by independent evaluation of SAHVAI performance, and then integration of **explainable AI (XAI)** features. Our project for detecting SAH and ICH with artificial intelligence **(SAHDAI)** comprises several phases.

The process began with a **manual detection phase**, during which potential hemorrhagic areas within cisternal spaces were identified. Each image slice was manually segmented, and any suspected bleeding regions were highlighted. In the next step, each image was annotated as either positive or negative for bleeding in an Excel sheet. These annotations were based on previously completed datasets, created through the combined efforts of clinical experts, including neuroradiologists, neurologists, and neurosurgeons. The annotated images were then uploaded to the **Google Cloud Platform (GCP)** (Figure 6a).

**Figure 6:**
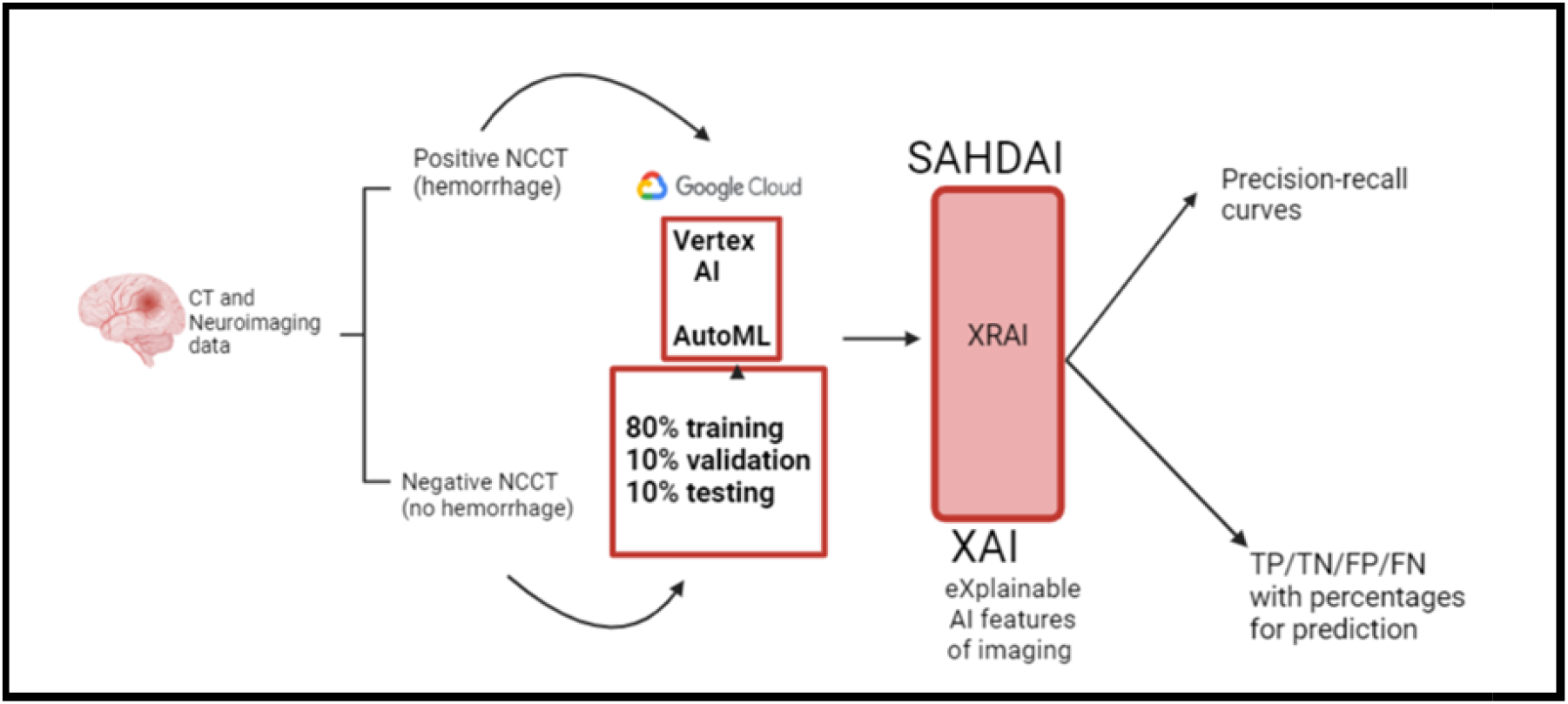
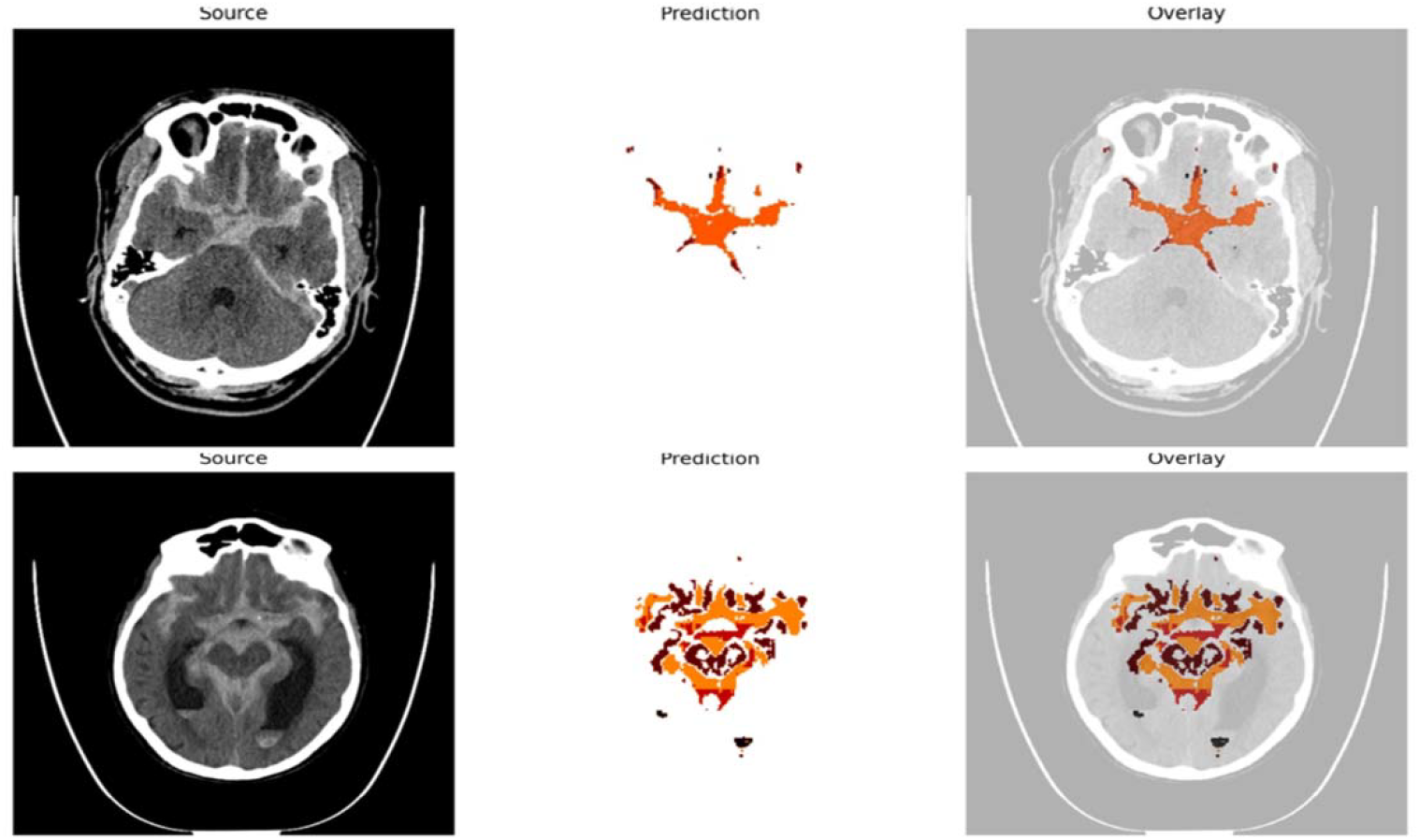
a) SAHDAI-XAI workflow explanation. **SAHDAI**-SAH Detection AI, **XAI**-eXplainable AI b) **Explainability and Data Interpretation:** The outcome percentages on GCP’s Vertex AI AutoML are presented, showing high and low true positives and true negatives based on the clarity and extent of the bleeding in the CT image. This enhances the model’s explainability. High true positive prediction (99.8%) Low true negative prediction (55.9%). The prediction outcomes were designed to mask the source images, demonstrating an overlay to enhance the visual explainability of the outcomes

On GCP, a machine learning model was developed and trained using **AutoML**, which automatically selected the optimal training settings to differentiate between slices with and without bleeding. The **XRAI** option was chosen to provide post-training explainability, generating heatmaps that highlight the regions most influential to the model’s decision-making.

The ML model processes images at a resolution of **256 × 3 RGB values per pixel**, with each pixel represented by three values (red, green, and blue). This results in approximately **16.77 million shades per pixel**, enabling the AI system to detect and analyze structures that may be imperceptible to the human eye (Figure 6b).

Ultimately, SAHDAI enables the discovery of novel phenotypes and biological patterns, addressing the fundamental question: *“Why is this happening?”*

### Integrating the UTS Framework: Understand Transform Sustain

The UTS (Understand, Transform, Sustain) framework, recently developed at Mayo Clinic, provides a structured yet simplified approach for integrating AI and process automation into healthcare. Rooted in quality improvement (QI) principles and continuous evaluation, UTS emphasizes sustained value and adaptability across projects. It serves as a pragmatic translation of broader frameworks, such as the Consolidated Framework for Implementation Research (CFIR), the Knowledge-to-Action (KTA) framework, and sustainment models in implementation science, into a streamlined structure aligned with the unique challenges of AI in healthcare (26–28). It is structured around three core phases: UNDERSTAND, TRANSFORM, and SUSTAIN. By bridging these established approaches with concepts from the learning healthcare system and the Total Product Life Cycle (TPLC) for AI-enabled devices, UTS provides a context-specific roadmap that connects QI tools with responsible and scalable AI implementation (29,30).

UTS provides the scaffolding for deploying SAHVAI and HEADS-UP as part of the CODE-ICH program, ensuring that their implementation is not only technically robust but also ethically sound, stakeholder-driven, and sustainable over time.

## Methods

### Study Design and Data Sources

This study used a mixed retrospective–prospective design. Non-contrast CT (NCCT) head scans from patients with spontaneous intracerebral and subarachnoid hemorrhage were obtained from the Mayo Clinic Comprehensive Stroke Center and affiliated hospitals (2015– 2024). The dataset included approximately *N* ≈*1000 CT* studies, covering diverse patient demographics and varying hemorrhage severities. Institutional Review Board approval was obtained with waiver of informed consent due to the de-identified nature of the data.

### Annotation and Ground Truth Generation

CT slices were reviewed and annotated independently by three board-certified neuroradiologists and two neurosurgeons. Hemorrhagic regions in cisternal and parenchymal compartments were manually segmented using ITK-SNAP software. Disagreements were resolved by consensus. An Excel-based labeling sheet categorized slices as positive or negative for bleeding, forming the ground truth for training and validation. Inter-rater reliability was measured with Cohen’s κ (target >0.85).

### Model Development

- **HEADS-UP:** DICOM images from emergency departments were uploaded to Google Cloud Platform (GCP) and processed through a custom AutoML pipeline for binary classification (hemorrhage vs no hemorrhage). Images were downsampled to 256 × 256 resolution and normalized. Data augmentation (rotation, intensity scaling, flipping) was applied to reduce overfitting.
- **SAHVAI:** A convolutional neural network (CNN) based on a U-Net backbone was trained to segment subarachnoid blood volumes. The model was optimized using Dice loss and Adam optimizer with a learning rate of 1e-4, batch size of 8, and early stopping criteria. Training was performed for up to 200 epochs.
- **Explainability (SAHDAI):** The XRAI overlay method was enabled to generate pixel-level heatmaps, ensuring transparency of predictions and identifying regions most influential in classification.

### Validation and Evaluation

Performance was assessed on an independent test cohort of *N* ≈ *1000* patients. Metrics included Dice similarity coefficient, accuracy, sensitivity, specificity, and AUC. SAHVAI outputs were compared against manual volumetric measurements, with Bland–Altman plots and intraclass correlation coefficients (ICC) used to confirm reproducibility. SAHV >10 mL at baseline was predefined as a clinically significant threshold for poor outcome prediction.

### Clinical Integration and Workflow Testing

The “ICH Phases” paging system was launched at Mayo Clinic CSC in December 2022. HEADS-UP was integrated for Phase I detection: ED CT scans were transmitted to the cloud, analyzed in under 10 seconds, and results returned to the stroke team via secure notification. SAHVAI outputs were embedded into PACS/EMR, providing 2D/3D/4D volumetric maps and real-time alerts for volumes exceeding 10 mL.

### Statistical Analysis

All analyses were conducted using Python (TensorFlow, PyTorch) and R (v4.3). Significance was defined at p < 0.05. Continuous variables were expressed as mean ± SD, categorical variables as percentages. Sensitivity analyses explored performance across subgroups (e.g., lobar vs deep hemorrhage, different scanners).

### UNDERSTAND: Building the Foundation

This phase focuses on establishing a robust foundation of trust, data integrity, and stakeholder alignment before the implementation of AI solutions.

- **Problem Framing & Clinical Context:** Hemorrhagic strokes, with a 30-day mortality rate of approximately 40%, critically lack time-sensitive protocols seen in ischemic strokes. The CODE-ICH framework addresses this by redefining ICH as an ultra-emergency, aligning with the “time is brain” principle.
- **Data Quality & Representation:** The SAHVAI and HEADS-UP systems are built upon Non-Contrast CT (NCCT) imaging and Electronic Medical Record (EMR) data, including intracranial pressure (ICP), cerebral perfusion pressure (CPP), electroencephalography (EEG), and transcranial doppler ultrasound (TCD). A strong emphasis is placed on utilizing diverse datasets to mitigate bias in hemorrhage detection and prediction.
- **Bias & Equity Considerations:** The project actively addresses design framing bias, data bias, and systemic bias. SAHDAI (Explainable AI) incorporates XAI overlays to visually demonstrate model reasoning, ensuring transparency and promoting equitable outcomes.
- **Stakeholder Engagement:** Success hinges on multidisciplinary collaboration, involving neurointensivists, radiologists, IT professionals, informaticians, and frontline clinicians. “Team Cards” will be utilized to document assumptions, acknowledge positionality, and address unconscious bias.
- **Regulatory & Ethical Readiness:** The framework aligns with established regulatory guidelines, including FDA’s SaMD (Software as a Medical Device) policies and IMDRF (International Medical Device Regulators Forum) frameworks. It incorporates principles of transparency, accountability, and explainability consistent with FDA’s Good Machine Learning Practice (GMLP) and anticipated EU AI Act requirements for high-risk medical AI. It also addresses GDPR Article 22, ensuring transparency in automated decision-making processes.

### TRANSFORM: Implementing and Iterating Solutions

This phase focuses on the practical implementation and iterative refinement of AI solutions, leveraging quality improvement (QI), change management, and implementation science tools.

- **Root Cause Analysis & QI Tools:** Fishbone diagrams and Root Cause Analysis (RCA) are employed to pinpoint delays in ICH care, such as those related to imaging interpretation and transfer times. Plan-Do-Study-Act (PDSA) cycles are applied to pilot SAHVAI and HEADS-UP within real-time Emergency Department (ED), Neuro-IR and Neurocritical Care Unit (NICU) workflows.
- **Agile Implementation:** SAHVAI seamlessly integrates 2D, 3D, and 4D volumetric analysis into NICU workflows. HEADS-UP utilizes cloud-based AI to detect ICH and trigger early alerts, thereby significantly reducing time-to-intervention.
- **Change Management (ADKAR):** The ADKAR model guides change management:
  - **Awareness:** Using the example of Sepsis bundle as a reference, ICH should be framed as urgent medical emergencies.
  - **Desire:** Clinician buy-in is fostered through real-time dashboards and explainable AI outputs. Our research, building on the work of Dr. Sakusic et al.’s RISE-HD (Abstract to be referenced) and utilizing 10 years of Florida FIOD data from the Kern Center for the Science of Healthcare Delivery, clearly demonstrates that hemorrhagic stroke is at least four times deadlier than ischemic stroke. Despite this stark difference, hemorrhagic stroke has historically received less attention due to its lower prevalence compared to ischemic stroke—a trend we are now actively working to reverse.
  - **Knowledge:** Comprehensive training is provided on AI interpretation and workflow integration.
  - **Ability:** AI tools are embedded directly into Picture Archiving and Communication Systems (PACS) and Electronic Medical Records (EMR).
  - **Reinforcement:** Continuous feedback loops and performance dashboards ensure sustained adoption.
- **Clinical Validation:** SAHVAI has been rigorously validated against the gold standard of manual segmentation. It demonstrates high predictive accuracy for Delayed Cerebral Ischemia (DCI) and poor outcomes, with SAHV greater than 10 mL tripling the risk. The system is already deployed in real-world settings through Mayo Clinic’s “ICH Phases” paging system.

### SUSTAIN: Ensuring Long-Term Reliability and Trust

This final phase focuses on maintaining the long-term reliability, scalability, and trustworthiness of the AI systems. *show projections of impact in populations…adpt scenarios 20,40,80%.. Economic savings and more lives saved.

- **Monitoring & Maintenance:** SAHVAI incorporates automated dashboards and control charts for continuous performance tracking. Plans are in place for continuous retraining and recalibration to address potential model drift.
- **Knowledge Management:** Lessons learned from deployment, such as the underutilization of pause features or resistance to AI, are meticulously documented and shared. Quality tools, including A3 reports, control plans, and transition plans, can be utilized.
- **Ethical Stewardship:** SAHDAI ensures explainability and fairness through XRAI overlays and transparent model operation. Continuous audits and stakeholder feedback loops are integral to the sustainment plan.
- **Scalability & Diffusion:** HEADS-UP addresses geographic disparities by enabling early detection in underserved areas. SAHVAI’s modular design facilitates seamless integration across various stroke centers and NICUs throughout the enterprise.

**Table.** Proposed AI implementation structured around three core phases: UNDERSTAND, TRANSFORM, and SUSTAIN to The CODE-ICH.

|  |  |
| --- | --- |
| UTS Phase | CODE-ICH Application |
| Understand | Data quality, bias mitigation, stakeholder alignment, regulatory readiness |
| Transform | Agile pilots, PDSA cycles, ADKAR change management, clinical validation |
| Sustain | Dashboards, control plans, explainability (SAHDAI), scalability (HEADS-UP, SAHVAI) |

### Broader relevance, challenges, and future directions

The clinical implications of HEADS-UP and SAHVAI are profound. Incorporating rapid detection and volumetric analysis directly into hospital PACS and EMR workflows reduces subjectivity, eliminates discrepancies in decision-making, and improves outcomes, particularly in areas with a shortage of healthcare personnel. Unlike the modified Fisher scale (mFS), which is subjective, SAHVAI provides objective and reproducible measurements. Consistent with Hemphill et al., SAHVAI augments rather than replaces clinical expertise, relieving neurointensivists of repetitive tasks and enabling them to focus on individualized decision-making (11). Such augmentation is increasingly necessary in the face of workforce shortages and the expanding demands of the NICU.

Evidence from Parry-Jones et al. and the INTERACT-3 trial demonstrates that bundled interventions improve outcomes. CODE-ICH adopts this philosophy in the management of ICH, unifying rapid diagnosis, volumetric analysis, and predictive modeling into a single workflow.

Implementation and Validation: Successful adoption of SAHVAI requires regulatory approval, transparency, and rigorous external validation to ensure reproducibility across institutions. Algorithms must be explainable and shareable, while avoiding automation bias through continued clinician engagement. Ethical considerations must remain at the forefront of deployment.

The next steps begin with full integration of SAHVAI into Mayo Clinic’s PACS system for initial validation. This phase will involve training clinicians on the SAHVAI workflow for real-time SAH volume measurement, benchmarking baseline accuracy and processing speed against manual methods, and collecting user feedback on speed, usability, and clinical impact.

Subsequent aims include implementing SAHVAI’s probabilistic risk prediction model in critical care units, with training to ensure use in at least 80% of eligible cases. A performance threshold of 85% sensitivity and specificity for predicting DCI will be established. Initial outcome metrics will include a 10% reduction in time-to-intervention for high-risk DCI patients.

Expansion and Deployment: SAHVAI will then be extended to Mayo Clinic’s Telestroke network, implemented across all 30 sites. The goal is to standardize volumetric output, conduct a multicenter case series demonstrating early risk detection, and test the hypothesis that SAHVAI improves intervention times by at least 15% compared with pre-implementation workflows. Regular usage by over 90% of clinicians across Telestroke sites will establish SAHVAI as a standard assessment tool.

Full deployment across all Mayo Clinic sites is planned, integrating SAHVAI into standard workflows for critical care and neurology units. Early outcome data will be collected on SAHVAI’s clinical impact, with targets including a 20% reduction in time-to-intervention for SAH patients. Interim results on accuracy and clinician satisfaction will be published.

International expansion will follow, with SAHVAI deployed in at least three global partner institutions across both high- and low-resource settings. Training programs will prepare 50+ clinicians worldwide, with the goal of achieving 90% accuracy in SAH volume assessment and DCI prediction. Baseline data will be collected to assess effectiveness and guide adjustments for varying healthcare environments.

Regulatory and Clinical Validation: SAHVAI will undergo regulatory submission, supported by data demonstrating at least a 15% improvement in DCI detection accuracy compared with conventional methods. A comprehensive analysis of clinical outcomes—including survival rates and ICU length of stay—will be published in a leading peer-reviewed journal.

Future Directions: The final stage involves conducting a large-scale, prospective international study evaluating mortality, ICU stay duration, and DCI incidence. By 2027, SAHVAI aims to expand to an additional five international sites, establishing global scalability.

Ultimately, the vision for HEADS-UP and SAHVAI extends beyond immediate applications. Both systems are designed to evolve into components of a continuous learning health system, refining performance with every patient encounter. The long-term goal is to integrate Tele–NeuroICU capabilities with real-time AI, transitioning the NICU from a reactive model to a predictive one—transforming the mantra from “time is brain” to “AI saves brain.”

## Conclusion

The integration of CODE-ICH, HEADS-UP, and SAHVAI represents a paradigm shift in the management of ICH. By embedding AI-driven detection, volumetric quantification, and predictive analytics into real-time clinical workflows, these tools address longstanding challenges in neurocritical care—namely, diagnostic delays, subjective assessments, and workforce limitations.

Unlike traditional scales such as the modified Fisher scale, SAHVAI provides objective, reproducible, and clinically actionable outputs within seconds, enabling timely interventions during the critical “golden hour.” HEADS-UP extends this capability to underserved populations through cloud-based triage and early alerting, bridging geographic and resource gaps.

Importantly, the SAHDAI framework introduces explainability into AI imaging, fostering clinician trust and regulatory readiness. Together, these innovations shift the NICU from a reactive to a predictive model of care, aligning with the evolving vision of a learning health system.

This transformation is best understood through the lens of the UTS framework. The Understand phase involves identifying gaps in hemorrhagic stroke care and leveraging high-fidelity data to build equitable AI models. The Transform phase operationalizes these models into clinical workflows, enabling real-time decision support and multidisciplinary coordination. Finally, the Sustain phase ensures long-term reliability through continuous monitoring, explainability, and integration into PACS and EMR systems. This framework supports not only technological innovation but also cultural and operational change across neurocritical care environments.

Looking ahead, the future of hemorrhagic stroke care lies in the convergence of tele-neurocritical care, real-time AI, and continuous model refinement. This work lays the foundation for a new clinical paradigm—one where “AI saves brain” becomes the operational mantra, and precision neurocritical care is accessible, scalable, and equitable.

## Data Availability

All data produced in the present study are available upon reasonable request to the authors

